# The cognitive profile of the schizophrenia-associated 3q29 deletion syndrome and the reciprocal 3q29 duplication syndrome

**DOI:** 10.64898/2026.11.05.26367369

**Authors:** Rebecca M Pollak, Daniela V Bishop, Matthew K Harner, Ariana Khan, John R Purcell, Madeleine Wheeler, Teresa Irving, Tyler M Moore, Kosha Ruparel, Ruben C Gur, Raquel E Gur, Jennifer G Mulle

**Affiliations:** Department of Psychiatry, Robert Wood Johnson Medical School, Rutgers University; Center for Advanced Biotechnology and Medicine, Rutgers University; Department of Psychiatry, Perelman School of Medicine, University of Pennsylvania; CHOP-Penn Lifespan Brain Institute, University of Pennsylvania

**Author notes:** Corresponding author information: Rebecca M Pollak, PhD; Center for Advanced Biotechnology and Medicine and Department of Psychiatry, Robert Wood Johnson Medical School, Rutgers University, 679 Hoes Lane West, Piscataway, NJ 08854. Ethics approval: This study was approved by Rutgers University’s Institutional Review Board (PRO2021001976). Consent to participate: All study subjects gave informed consent prior to participating in this study. Consent to publish: Not applicable. Welfare of animals: Not applicable. Availability of data and materials: The datasets used and/or analyzed during the current study are available from the corresponding author on reasonable request.

## Abstract

3q29 deletion syndrome (3q29del) and the reciprocal 3q29 duplication syndrome (3q29dup) are associated with a range of neuropsychiatric phenotypes, including impaired cognition. However, previous sample sizes have been small, and cognitive performance has not been systematically compared between 3q29del and 3q29dup. For both disorders, the impact of familial background on cognition is unknown. 3q29del is associated with high risk for schizophrenia; in the general population individuals at Clinical High Risk (CHR) for psychosis exhibit impaired cognition, but the relationship between CHR status and cognition in 3q29del is unexplored. Here we address these questions in the largest sample of 3q29del and 3q29dup ever evaluated. Individuals with 3q29del (n=103, 58.3% male), 3q29dup (n=38, 55.3% male), and controls (n=54, 42.6% male) were assessed with the Peabody Picture Vocabulary Test, 5^th^ Edition (PPVT-5) and the Penn Computerized Neurocognitive Battery (Penn-CNB). Total PPVT-5 scores and global, domain-level, and task-level Penn-CNB scores were compared across groups. Compared to controls, 3q29del participants scored an average of 24.6 and 22.9 points lower on the PPVT and Penn CNB, respectively (p<3E-16); 3q29dup participants scored an average of 18.8 and 14.2 points lower on the PPVT and Penn CNB (p<4E-6). In addition, 3q29del and 3q29dup exhibit unique profiles of cognitive deficits. To assess the impact of familial background on cognition, we evaluated unaffected relatives of 3q29del (n=140, 38.6% male), 3q29dup (n=25, 24.0% male), and controls (n=45, 11.1% male). The average familial shift on the Penn-CNB was 22.4 points for 3q29del and 16.7 points for 3q29 dup. Individuals with 3q29del+CHR exhibited greater cognitive deficits than 3q29del-CHR. These results demonstrate that both 3q29del and 3q29dup are associated with significant cognitive impairment and indicate that cognitive testing should be standard of care for individuals diagnosed with 3q29del or 3q29dup.

## Introduction

Reciprocal copy number variants (CNVs) in the 3q29 region at the end of the long arm of chromosome 3 are associated with significant neurodevelopmental and psychiatric pathology. The region is the site of two CNV syndromes, 3q29 deletion syndrome (3q29del) and 3q29 duplication syndrome (3q29dup) (hg19, chr3:195725000-197350000). 3q29del is a rare (∼1:30,000) typically *de novo* deletion of the 3q29 interval (1, 2, 3, 4, 5), and is characterized by a greater than 40-fold increased risk for schizophrenia (6, 7, 8, 9, 10), a 19-fold increased risk for autism spectrum disorder (ASD) (11, 12, 13), and increased risk for mild to moderate intellectual disability (ID), anxiety disorders, and attention deficit/hyperactivity disorder (ADHD) (3, 4, 5, 14, 15, 16, 17). The reciprocal 3q29dup is also rare, with prevalence estimates in the general population ranging from ∼1:75,000 to ∼1:8,000 (1, 18, 19, 20, 21), and has been reported in the literature as both *de novo* and inherited cases (4, 22, 23, 24). Individuals with 3q29dup are at increased risk for developmental delay, ID, obesity, seizures, disruptive behavior, and ASD (4, 22, 23, 24, 25, 26, 27, 28, 29, 30, 31, 32). While there is some phenotypic overlap between 3q29del and 3q29dup, including increased risk for ID, cognitive ability in these disorders has never been directly compared.

While both 3q29del and 3q29dup are associated with a substantially increased risk for a range of neurodevelopmental and psychiatric phenotypes, there is a significant degree of phenotypic heterogeneity between affected individuals. Some individuals with 3q29del or 3q29dup are able to live fully independent lives with cognitive and social function indistinguishable from typically developing controls, whereas other individuals with 3q29del or 3q29dup have severe phenotypes and may require lifelong care. This phenomenon is not unique to the 3q29 CNVs; phenotypically similar CNVs such as 22q11.2 deletion and duplication syndromes and 16p11.2 deletion and duplication syndromes are also characterized by phenotypic heterogeneity (14, 33, 34, 35, 36, 37, 38, 39, 40, 41, 42, 43, 44, 45, 46). Some of this phenotypic heterogeneity may be explained by familial background effects, whereby the familial genetic background influences the degree of expression of CNV-related phenotypes (47). Indeed, familial background effects have been reported in the relationship between the 22q11.2 deletion and cognitive ability (48, 49), and in the relationship between both the 16p11.2 deletion and duplication and cognitive ability, social function, and body mass index (50, 51, 52). The effect of parental phenotype and familial background has not been explored in 3q29del or 3q29dup.

We have previously developed an optimized, validated **remote** phenotyping protocol that allowed us to collect phenotype data, including cognitive ability, on study participants with rare genomic syndromes at scale (53). In the present study we conducted remote phenotyping on over 400 study participants to address the following questions: what is the relative impact of 3q29del and 3q29dup on cognitive function, and what is the impact of familial background on cognitive phenotypes in 3q29del and 3q29dup? Prior publications on 3q29del are case reports or small sample sizes (3, 15, 17); a previous report in 3q29dup lacked systematic collection of cognitive ability (24). To address these issues, we recruited the largest cohort of systematically evaluated individuals with 3q29del and 3q29dup to date: we evaluated 103 3q29del carriers and 38 3q29dup carriers. To explore the influence of familial background on cognitive phenotypes, we also evaluated 140 unaffected relatives of 3q29del probands and 25 unaffected relatives of 3q29dup probands. We included 54 typically developing controls and 45 relatives of controls as a general population comparison group. The results from this study will improve our current understanding of the 3q29del and 3q29dup cognitive phenotype, as well as provide the first analysis of the role of familial background on the expression of syndromic phenotypes.

## Methods

### Study Participants

Individuals with 3q29del (n=103, 58.25% male, 16.98±11.17 years old) or 3q29dup (n=38, 55.26% male, 20.34±13.67 years old) were recruited from the existing online 3q29 registry (3q29deletion.com) (5), as well as direct referrals and contacts. Individuals with a 3q29 deletion or duplication residing completely outside of the canonical 1.6 Mb interval were excluded from analysis. Unaffected relatives of probands with 3q29del (n=140, 38.57% male, 46.86±8.79 years old) or 3q29dup (n=25, 24.00% male, 51.40±10.96 years old) were also enrolled. Typically developing controls (n=54, 42.59% male, 17.85±10.66 years old) and their relatives (n=45, 11.11% male, 46.09±8.16 years old) were recruited from community sources and from ResearchMatch, an online platform for individuals interested in participating in research (54). A description of the study sample can be found in Table 1 and Figure 1. This study was approved by Rutgers University’s Institutional Review Board (PRO2021001360).

**Figure 1.**
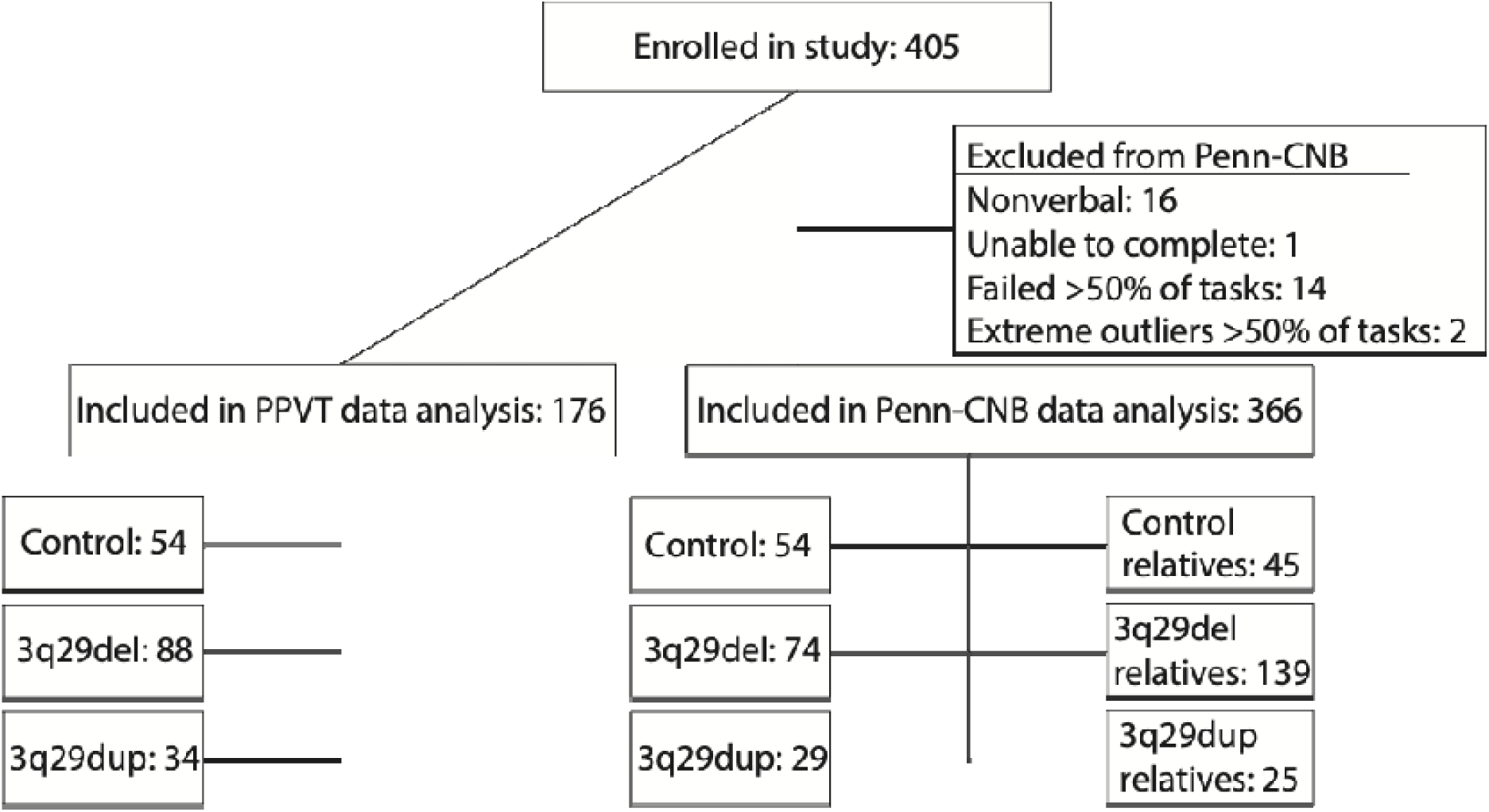
Consort diagram showing total study enrollment, participants excluded from Penn-CNB analysis, and final sample included in analysis across all groups.

**Table 1.** Demographics of study participants.

|  |  |  | Total enrollment | Completed PPVT-5 | Completed Penn-CNB | Completed PPVT and Penn-CNB |
|---|---|---|---|---|---|---|
| Control | N |  | 54 | 54 | 54 | 54 |
|  | Age (years) |  |  |  |  |  |
|  |  | Mean ± SD | 17.85 ± 10.66 | 17.85 ± 10.66 | 17.85 ± 10.66 | 17.85 ± 10.66 |
|  |  | Range | 8 - 62 | 8 - 62 | 8 - 62 | 8 - 62 |
|  | Sex |  |  |  |  |  |
|  |  | Male | 23 (42.59%) | 23 (42.59%) | 23 (42.59%) | 23 (42.59%) |
|  |  | Female | 31 (5741%) | 31 (5741%) | 31 (5741%) | 31 (5741%) |
|  | Race |  |  |  |  |  |
|  |  | White | 45 (83.33%) | 45 (83.33%) | 45 (83.33%) | 45 (83.33%) |
|  |  | Other | 9 (16.67%) | 9 (16.67%) | 9 (16.67%) | 9 (16.67%) |
|  | Ethnicity |  |  |  |  |  |
|  |  | Hispanic/Latino | 6 (11.11%) | 6 (11.11%) | 6 (11.11%) | 6 (11.11%) |
|  |  | Not Hispanic/Latino | 48 (88.89%) | 48 (88.89%) | 48 (88.89%) | 48 (88.89%) |
| 3q29 deletion | N |  | 103 | 88 | 100 | 85 |
|  | Age (years) |  |  |  |  |  |
|  |  | Mean ± SD | 16.98 ± 11.17 | 17.57 ± 11.78 | 16.72 ± 10.94 | 17.28 ± 11.56 |
|  |  | Range | 7 - 68 | 7 - 68 | 7 - 68 | 7 - 68 |
|  | Sex |  |  |  |  |  |
|  |  | Male | 60 (58.25%) | 49 (55.68%) | 57 (57.00%) | 46 (54.12%) |
|  |  | Female | 43 (41.75%) | 39 (44.32%) | 43 (43.00%) | 39 (45.88%) |
|  | Race |  |  |  |  |  |
|  |  | White | 96 (93.20%) | 82 (93.18%) | 93 (93.00%) | 79 (92.94%) |
|  |  | Other | 7 (6.80%) | 6 (6.82%) | 7 (7.00%) | 6 (7.06%) |
|  | Ethnicity |  |  |  |  |  |
|  |  | Hispanic/Latino | 9 (8.74%) | 8 (9.09%) | 8 (8.00%) | 7 (8.24%) |
|  |  | Not Hispanic/Latino | 94 (91.26%) | 80 (90.91%) | 92 (92.00%) | 78 (91.76%) |
| 3q29 duplication | N |  | 38 | 34 | 35 | 31 |
|  | Age (years) |  |  |  |  |  |
|  |  | Mean ± SD | 20.34 ± 13.67 | 21.56 ± 13.91 | 19.74 ± 13.26 | 21.00 ± 13.54 |
|  |  | Range | 6 - 48 | 6 - 48 | 7 - 48 | 7 - 48 |
|  | Sex |  |  |  |  |  |
|  |  | Male | 21 (55.26%) | 19 (55.88%) | 18 (51.43%) | 16 (51.61%) |
|  |  | Female | 17 (44.74%) | 15 (44.12%) | 17 (48.57%) | 15 (48.39%) |
|  | Race |  |  |  |  |  |
|  |  | White | 33 (86.84%) | 30 (88.24%) | 30 (85.71%) | 27 (87.10%) |
|  |  | Other | 5 (13.16%) | 4 (11.76% | 5 (14.29%) | 4 (12.90%) |
|  | Ethnicity |  |  |  |  |  |
|  |  | Hispanic/Latino | 3 (7.89%) | 2 (5.88%) | 3 (8.57%) | 2 (6.45%) |
|  |  | Not Hispanic/Latino | 35 (92.11%) | 32 (94.12%) | 32 (91.43%) | 29 (93.55%) |
| Control relatives | N |  | 45 |  | 45 |  |
|  | Age (years) |  |  |  |  |  |
|  |  | Mean ± SD | 46.09 ± 8.16 |  | 46.09 ± 8.16 |  |
|  |  | Range | 28 - 74 |  | 28 - 74 |  |
|  | Sex |  |  |  |  |  |
|  |  | Male | 5 (11.11%) |  | 5 (11.11%) |  |
|  |  | Female | 40 (88.89%) |  | 40 (88.89%) |  |
|  | Race |  |  |  |  |  |
|  |  | White | 37 (82.22%) |  | 37 (82.22%) |  |
|  |  | Other | 8 (17.78%) |  | 8 (17.78%) |  |
|  | Ethnicity |  |  |  |  |  |
|  |  | Hispanic/Latino | 3 (6.67%) |  | 3 (6.67%) |  |
|  |  | Not Hispanic/Latino | 42 (93.33%) |  | 42 (93.33%) |  |
| 3q29 deletion relatives | N |  | 140 |  | 140 |  |
|  | Age (years) |  |  |  |  |  |
|  |  | Mean ± SD | 46.86 ± 8.79 |  | 46.86 ± 8.79 |  |
|  |  | Range | 31 - 72 |  | 31 - 72 |  |
|  | Sex |  |  |  |  |  |
|  |  | Male | 54 (38.57%) |  | 54 (38.57%) |  |
|  |  | Female | 86 (61.43%) |  | 86 (61.43%) |  |
|  | Race |  |  |  |  |  |
|  |  | White | 127 (90.71%) |  | 127 (90.71%) |  |
|  |  | Other | 13 (9.29%) |  | 13 (9.29%) |  |
|  | Ethnicity |  |  |  |  |  |
|  |  | Hispanic/Latino | 7 (5.00%) |  | 7 (5.00%) |  |
|  |  | Not Hispanic/Latino | 133 (95.00%) |  | 133 (95.00%) |  |
| 3q29 duplication relatives | N |  | 25 |  | 25 |  |
|  | Age (years) |  |  |  |  |  |
|  |  | Mean ± SD | 51.40 ± 10.96 |  | 51.40 ± 10.96 |  |
|  |  | Range | 37 - 80 |  | 37 - 80 |  |
|  | Sex |  |  |  |  |  |
|  |  | Male | 6 (24.00%) |  | 6 (24.00%) |  |
|  |  | Female | 19 (76.00%) |  | 19 (76.00%) |  |
|  | Race |  |  |  |  |  |
|  |  | White | 21 (84.00%) |  | 21 (84.00%) |  |
|  |  | Other | 4 (16.00%) |  | 4 (16.00%) |  |
|  | Ethnicity |  |  |  |  |  |
|  |  | Hispanic/Latino | 1 (4.00%) |  | 1 (4.00%) |  |
|  |  | Not Hispanic/Latino | 24 (96.00%) |  | 24 (96.00%) |  |

### Measures

The measures used in this study were as previously described (53). Briefly, receptive verbal comprehension was assessed using the Peabody Picture Vocabulary Test, Fifth Edition (PPVT-5) (55). The PPVT-5 requires the study participant to listen to a word and select one of four illustrations that best depicts the word. Neurocognitive ability was assessed using the Penn Computerized Neurocognitive Battery (Penn-CNB), measuring of cognitive ability across five domains: executive control, episodic memory, complex cognition, social cognition, and sensory/motor speed (56). The Penn-CNB comprises 13 distinct computerized tasks designed to measure the accuracy and speed of participants’ responses (full list of tasks in Supplemental Material). Both the PPVT and Penn-CNB were included based on prior work by our team demonstrating significant differences between verbal and non-verbal cognitive ability (16). To identify individuals with 3q29del at Clinical High Risk (CHR) for psychosis, the Structured Interview for Psychosis-Risk Syndromes (SIPS) (57) was used. The SIPS is a gold-standard semi-structured interview used to assess prodromal symptoms of psychosis. Participants are rated on a scale of 0-6 across positive, negative, disorganized, and general symptom domains, with scores of 3-5 on any positive symptom domain item indicating CHR, and a score of 6 on any positive symptom domain item indicating psychosis. All evaluations were administered via a HIPAA-compliant videoconferencing platform (Zoom) by a trained research assistant.

### Penn-CNB scoring

Quality control procedures for the Penn-CNB were implemented according to developer recommendations (56). Briefly, a subject’s completion and effort on each task is scored by the test administrator on a scale of C1-C3 and I1-I3 (Figure S1). For each task a participant completes from start to finish, they are assigned a score between C1-C3 for that task. C1 means that the participant completed the task with good effort and understanding throughout, C2 means the participant completed the task with good effort on at least 50% of the task items, and C3 means the participant completed the task with good effort on less than 50% of the task items. If a participant is unable to complete a task, they are assigned a score between I1-I3 for that task. I1 means that the participant did not complete the task but showed good effort on the items they completed, I2 means the participant did not complete the task and had good effort on at least 50% of the completed task items, and I3 means the participant did not complete the task and had good effort on less than 50% of the completed task items. Tasks with a score of C1 or C2 were retained in the present analysis, tasks with a score of C3, I1, I2, or I3 were considered failed and the subject’s data for that task was removed from the analysis. If a subject failed 7 or more tasks (more than 50% of the assessment) they were removed from the analysis. Tasks with efficiency z scores more than 10 standard deviations from the mean were considered extreme outliers and the subject’s data for that task was removed from the analysis. If a subject recorded extreme outliers on 7 or more tasks they were removed from the analysis. A summary of the participants excluded from Penn-CNB analysis can be found in Table S1. A consort diagram (Figure 1) shows the number of subjects excluded from Penn-CNB analysis based on quality control measures. Data were winsorized with a 1% threshold prior to analysis using the Winsorize function in the DescTools R package (58). Detailed data processing information can be found in the Supplemental Material.

Age-and sex-adjusted z scores were calculated for each task using norms available from the Penn-CNB developers. Participant accuracy and speed are recorded for each Penn-CNB task, excluding the Motor Praxis and Short Computerized Finger Tapping Test, which generate a score for Speed only. The accuracy score for each task corresponds to the number of items the participant completed correctly for the task. The speed score for each task corresponds to the median elapsed time for the participant to record a correct response for the task (response time in msec). Speed z-scores were multiplied by-1 so that they consistently reflect better performance with higher scores. Efficiency scores for each task were calculated as the average of accuracy and speed for that task. Global Accuracy, Speed, and Efficiency metrics were calculated as the average of the task-specific accuracy, speed, and efficiency values with no imputation for missing data. Penn-CNB z scores were transformed to a normal distribution with mean of 100 and standard deviation of 15 for ease of interpretation.

### Analysis

All analyses were performed in R version 4.2.2 (59). Standardized scores were used for all analyses. Because both the PPVT-5 and Penn-CNB standardized scores are adjusted for age and sex, age and sex were not included as covariates in any models. Statistical analysis was performed using simple linear models, two sample t tests, and Pearson correlation tests implemented using the stats R package (59). Sample variances were compared using F-tests implemented using the stats R package (59). For the familial shift analysis, if two parents or relatives completed the assessment, a biparental Penn-CNB Global Accuracy mean was calculated. If only one parent or relative completed the Penn-CNB, their score was used without further manipulation. The familial shift was calculated by subtracting the relative Global Accuracy score from the proband’s score; probands that score lower than their relative(s) receive a negative shift value, and probands that score higher than their relative(s) receive a positive shift value. One-sample t-tests implemented using the stats R package (59) were used to determine if the average shift value was different from zero. The analysis was performed using all available relatives and was repeated with only first-degree relatives to assess the impact of including non-first-degree relatives in the analysis. Data visualization was performed using the plotly (60) and ggplot2 (61) R packages.

## Results

### Cognitive performance on the PPVT and Penn-CNB

#### Cognitive performance in 3q29del

On the PPVT-5, participants with 3q29del scored an average of 24.6 points lower than typically developing controls (3q29del mean=87.28±14.47, control mean=111.93±15.37; p=2.80E-16; Figure 2A). On the Penn-CNB, participants with 3q29del scored an average of 20.8 points lower than typically developing controls on Global Efficiency (3q29del mean=83.96±15.46, control mean=104.83±8.01; p=1.26E-15; Figure 2B), 22.9 points lower on Global Accuracy (3q29del mean=84.11±14.29, control mean=107.02±8.73; p<2E-16; Figure 2C), and 12.7 points lower on Global Speed (3q29del mean=87.13±17.17, control mean=99.87±8.05; p=1.95E-6; Figure 2D). We also compared the variance of the score distributions to assess the degree of phenotypic heterogeneity in 3q29del. For the PPVT-5, there was no difference in the variance of the score distribution between 3q29del and controls (p=0.61); however, for the Penn-CNB, 3q29del had significantly larger variance in the score distributions relative to controls for Global Efficiency (p=1.61E-6), Global Accuracy (p=0.0002), and Global Speed (p=4.79E-8). These data suggest that there is increased variability in cognitive performance in 3q29del as compared to controls, indicating significant phenotypic heterogeneity.

**Figure 2.**
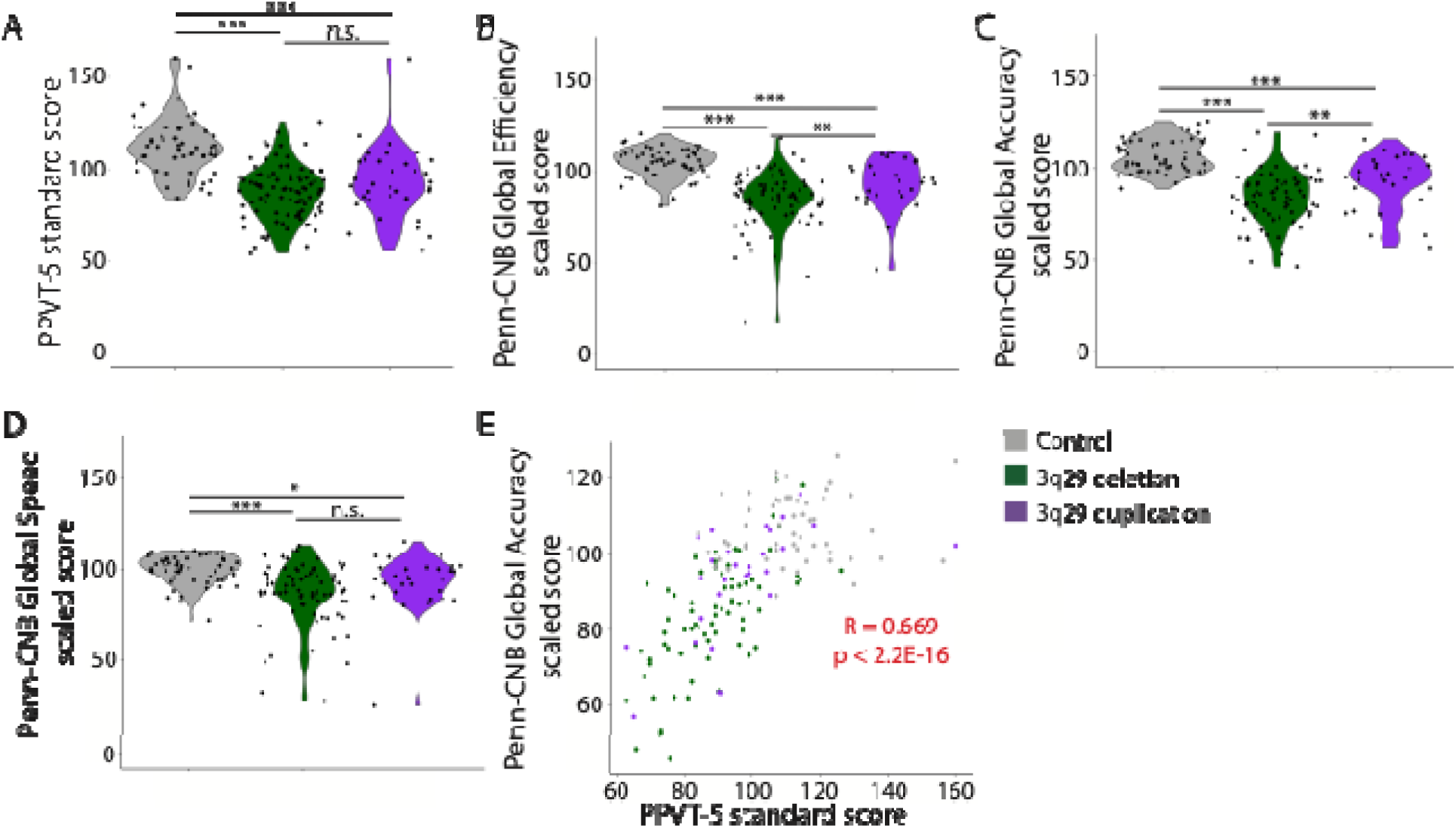
**A)** Distribution of scores on the PPVT-5 for controls (n=54), 3q29del (n=88), and 3q29dup (n=34). **B)** Distribution of Penn-CNB Global Efficiency scores for controls (n=54), 3q29del (n=74), and 3q29dup (n=29). **C)** Distribution of Penn-CNB Global Accuracy scores for controls (n=54), 3q29del (n=74), and 3q29dup (n=29). **D)** Distribution of Penn-CNB Global Speed scores for controls (n=54), 3q29del (n=74), and 3q29dup (n=29). **E)** Correlation between PPVT-5 scores and Penn-CNB Global Accuracy scores. n.s., not significant; *, p<0.05; **, p<0.01; ***, p<0.0001

#### Cognitive performance in 3q29dup

On the PPVT-5, participants with 3q29dup scored an average of 18.8 points lower than typically developing controls (3q29dup mean=93.12±19.17, control mean=111.93±15.37; p=1.65E-7; Figure 2A). On the Penn-CNB, participants with 3q29dup scored an average of 11.6 points lower than typically developing controls on Global Efficiency (3q29dup mean=93.19±13.94, control mean=104.83±8.01; p=0.0002; Figure 2B), 14.2 points lower on Global Accuracy (3q29dup mean=92.79±14.82, control mean=107.02±8.73; p=3.14E-6; Figure 2C), and 7.0 points lower for Global Speed (3q29dup mean=92.82±15.69, control mean=99.87±8.05; p=0.035; Figure 2D). We also compared the variance of the score distributions to assess the degree of phenotypic heterogeneity in 3q29dup. For the PPVT-5, there was no difference in the variance of the score distribution between 3q29dup and controls (p=0.149). For the Penn-CNB, 3q29dup had significantly larger variance in the score distributions relative to controls for Global Efficiency (p=0.0005), Global Accuracy (p=0.0009), and Global Speed (p=2.86E-5). These data show increased variability in cognitive performance in 3q29dup, similar to 3q29del.

#### Differences in cognitive performance between 3q29del and 3q29dup

Individuals with 3q29del and 3q29dup did not score significantly differently from each other on the PPVT-5 (p=0.12; Figure 2A). On the Penn-CNB, subjects with 3q29del scored on average 9.22 and 8.68 points lower than subjects with 3q29dup on Global Efficiency and Global Accuracy, respectively (Global Efficiency p=0.005, Global Accuracy p=0.009; Figure 2B-C), but not on Global Speed (p=0.11, Figure 2D), indicating that the 3q29 deletion has a greater impact on cognitive ability as measured by the Penn-CNB than the 3q29 duplication, and this is driven by differential performance on Global Accuracy.

#### The PPVT-5 and Penn-CNB measure distinct aspects of cognitive ability

Within our sample of 195 probands, we evaluated the correlation between the PPVT-5 and Penn-CNB to determine if they are measuring different aspects of cognition. We found a significant positive correlation between the PPVT-5 standard score and the Penn-CNB Global Accuracy score (R=0.669, p<2.2E-16; Figure 2E). The PPVT-5 standard score is significantly correlated with each Penn-CNB domain accuracy score, but the strength of the correlation varies. The strongest domain correlation is between the PPVT-5 standard score and the Complex cognition domain accuracy score (R=0.597, p=5.94E-16), and the weakest domain correlation is between the PPVT-5 standard score and the Executive domain accuracy scale (R=0.497, p=8.20E-11). Together, these data suggest that the PPVT-5 and the Penn-CNB are measuring related, but distinct, cognitive phenotypes in our study population.

### Domain-and task-specific deficits on the Penn-CNB

#### Penn-CNB domain-and task-specific deficits in 3q29del

We next analyzed Penn-CNB performance across domains to determine if the reduced Penn-CNB scores in 3q29del were due to specific domain challenges versus global deficits. We found that 3q29del participants had significant deficits compared to controls across all Penn-CNB domains for efficiency and accuracy (Figure 3A-B, Table S2, Table S3). Subjects with 3q29del had a 10-20% slower response speed compared to controls on four of the five Penn-CNB domains (Executive, Episodic memory, Social cognition, and Sensorimotor; all p<0.005), with a suggestive reduction in response speed on the Complex cognition domain (6% difference; p=0.078; Figure 3C, Table S4).

**Figure 3.**
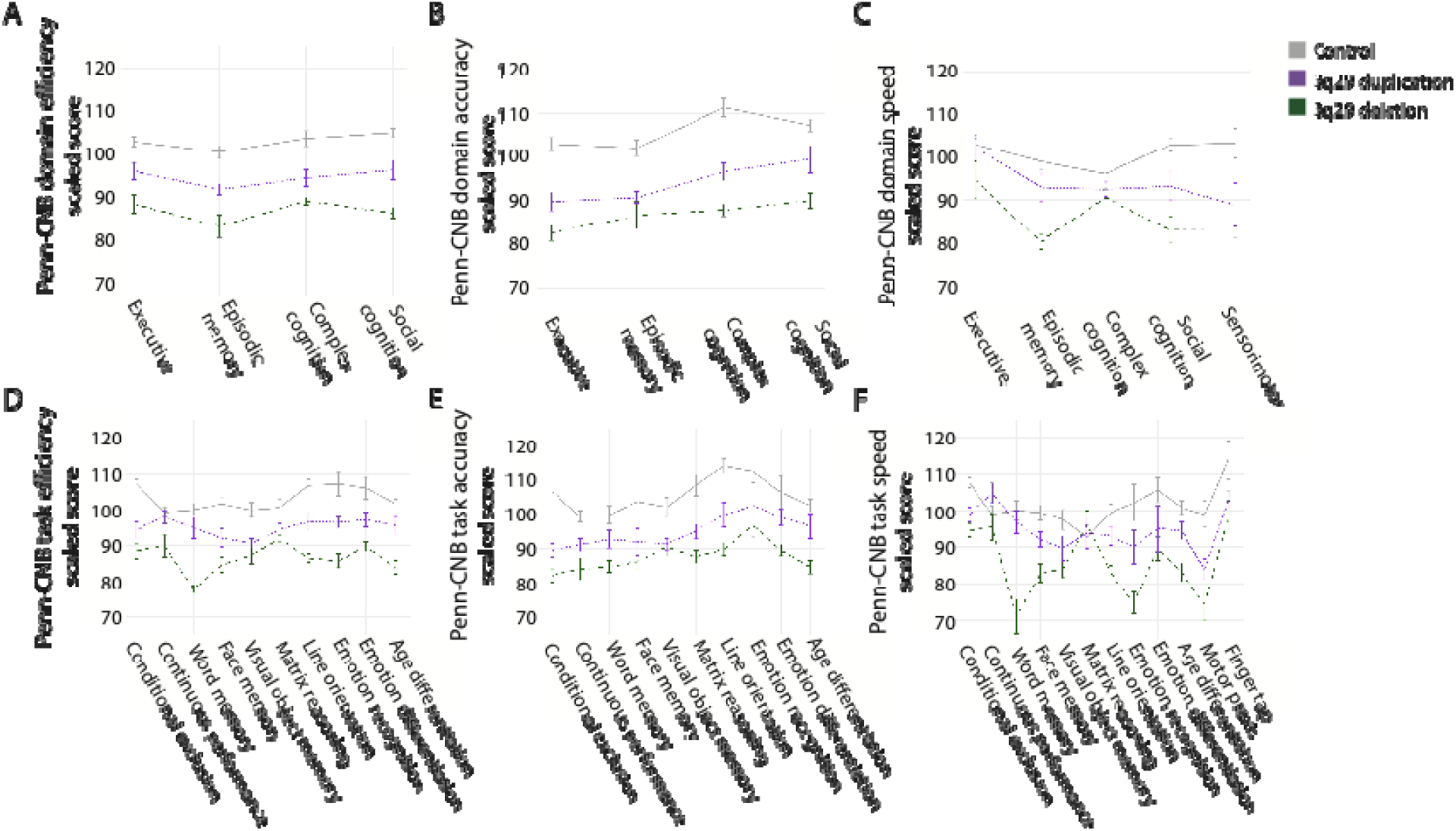
Penn-CNB domain-and task-levels scores for control (n=54), 3q29del (n=74), and 3q29dup (n=29). **A)** Domain efficiency scores. **B)** Domain accuracy scores. **C)** Domain speed scores. **D)** Task efficiency scores. **E)** Task accuracy scores. **F)** Task speed scores. Data are shown as mean ± SEM.

We further examined task-level data for subjects with 3q29del compared to controls. We found that individuals with 3q29del scored significantly lower on efficiency across all tasks relative to controls (Figure 3D, Table S2), with the most substantial deficit on the Word memory task (3q29del mean=77.93±28.08, control mean=100.04±12.60; p=1.11E-7). Separating accuracy from speed, individuals with 3q29del scored significantly lower on accuracy across all tasks relative to controls (Figure 3E, Table S3), with the most substantial deficit on the Conditional exclusion task (3q29del mean=82.41±15.01, control mean=106.83±14.16; p=5.14E-16). Individuals with 3q29del recorded significantly slower response speed than controls on ten of twelve tasks (Figure 3F, Table S4), with the most substantial deficit on the Word memory task (3q29del mean=70.97±43.63, control mean=100.10±17.60; p=2.10E-6). Response speed on the Continuous performance and Matrix reasoning tasks was relatively well-preserved in 3q29del participants compared to controls (p>0.300).

#### Penn-CNB domain-and task-specific deficits in 3q29dup

3q29dup participants had significant deficits compared to controls across all Penn-CNB domains for both efficiency and accuracy (Figure 3A-B, Table S2, Table S3). Individuals with 3q29dup had a 10-15% slower response speed compared to controls on two of the five domains (Social cognition and Sensorimotor; all p<0.05; Figure 3C, Table S4). There were no differences in response speed between 3q29dup subjects and controls for the Executive, Episodic memory, or Complex cognition domains (all p>0.250; Figure 3C, Table S4).

Analysis of task-level data revealed that individuals with 3q29dup scored significantly lower than controls on efficiency for seven of ten tasks (Figure 3D, Table S2), with the most substantial deficit on the Conditional exclusion task (3q29dup mean=94.25±10.25, control mean=107.28±9.59; p=3.46E-6). Individuals with 3q29dup scored significantly lower than controls on accuracy for six of ten tasks (Figure 3E, Table S3), with the most substantial deficit on the Conditional exclusion task (3q29dup mean=89.59±14.60, control mean=106.83±14.16; p=1.77E-6). Individuals with 3q29dup recorded significantly slower response speed than controls for four of twelve tasks (Figure 3F, Table S4), with the most substantial deficit on the Motor praxis task (3q29dup mean=84.00±34.34, control mean=99.03±19.10; p=0.045).

#### Distinct neurocognitive profiles between 3q29del and 3q29dup

We directly compared subjects with 3q29del and 3q29dup to determine if the degree of Penn-CNB deficit varied across domains between the syndromes. We found that for efficiency, subjects with 3q29del scored significantly lower than subjects with 3q29dup across all domains, with the most substantial difference in the Social cognition domain (3q29del mean=90.07±17.04, 3q29dup mean=99.46±16.86, p=0.014; Figure 3A, Table S2). Regarding accuracy, subjects with 3q29del scored significantly lower than subjects with 3q29dup on three of the four domains (Executive, Complex cognition, and Social cognition), with the most substantial difference in Social cognition (3q29del mean=87.93±14.38, 3q29dup mean=96.60±11.37, p=0.002), while accuracy on the Episodic memory domain was not significantly different between 3q29del and 3q29dup (p=0.267; Figure 3B, Table S3). Regarding speed, subjects with 3q29del were significantly slower than subjects with 3q29dup on three of the five speed domains (Executive, Episodic memory, and Social cognition), with the most substantial difference in Episodic memory (3q29del mean=80.07±30.50, 3q29dup mean=92.98±23.74, p=0.026), while response speed on the Complex cognition and Sensorimotor domains was not significantly different between 3q29del and 3q29dup (all p>0.300, Figure 3C, Table S4). Together, these data support a more severe cognitive deficit in individuals with 3q29del relative to 3q29dup, both in the degree of deficit and in the number of domains impacted.

We also compared task-level performance between study subjects with 3q29del and 3q29dup. Subjects with 3q29del had significantly worse efficiency scores than subjects with 3q29dup on seven of ten tasks (Figure 3D, Table S2), with the most substantial difference on the Word memory task (3q29del mean=77.93±28.08, 3q29dup mean=94.77±16.49; p=0.0006). Subjects with 3q29del and 3q29dup had similar efficiency scores on the Face memory, Visual object memory, and Matrix reasoning tasks (Figure 3D, Table S2). Subjects with 3q29del had significantly worse accuracy scores than subjects with 3q29dup on four of ten tasks (Figure 3E, Table S3), with the most substantial difference on the Age differentiation task (3q29del mean=84.79±20.78, 3q29dup mean=96.59±19.57; p=0.009). Subjects with 3q29del recorded significantly slower response speed than subjects with 3q29dup on five of twelve tasks (Figure 3F, Table S4), with the most substantial difference on the Word memory task (3q29del mean=70.97±43.63, 3q29dup mean=96.84±17.64; p=0.0001). Together, these data demonstrate wide-ranging cognitive deficits in both 3q29del and 3q29dup and highlight specific dimensions of cognition with more severe deficits in 3q29del.

### Effect of genetic background on Penn-CNB performance

#### Population-level differences in Penn-CNB performance

In the general population, cognitive ability is highly heritable (62, 63). To determine the influence of familial background on Penn-CNB performance, we evaluated unaffected relatives of controls, 3q29del probands, and 3q29dup probands. The Penn-CNB Global Accuracy score is the best proxy for IQ (64) and was the focus of this analysis. We first compared the distribution of proband Global Accuracy scores as compared to their respective relative population. There was no difference in Penn-CNB Global Accuracy score between controls and control relatives (control mean=107.02±8.73, control relative mean=107.60±6.51, p=0.706; Figure 4A). Subjects with 3q29del scored an average of 21.7 points, or 1.5 standard deviations, lower than their unaffected relatives (3q29del mean=84.11±14.29, 3q29del relative mean=105.76±8.58, p<2.2E-16). For 3q29dup, this was less pronounced with a mean difference of 12.9 points between probands and unaffected relatives (3q29dup mean=92.79±14.82, 3q29dup relative mean=105.66±9.09, p=0.0003; Figure 4A). We also compared the variances of the proband and relative distributions within each group: control, 3q29del, and 3q29dup proband distributions all had significantly greater variance compared to their respective unaffected family member distributions (control p=0.048, 3q29del p=2.43E-7, 3q29dup p=0.016; Figure 4A). Together, these data demonstrate a significant cognitive deficit in 3q29del and 3q29dup probands compared to their unaffected relatives.

**Figure 4.**
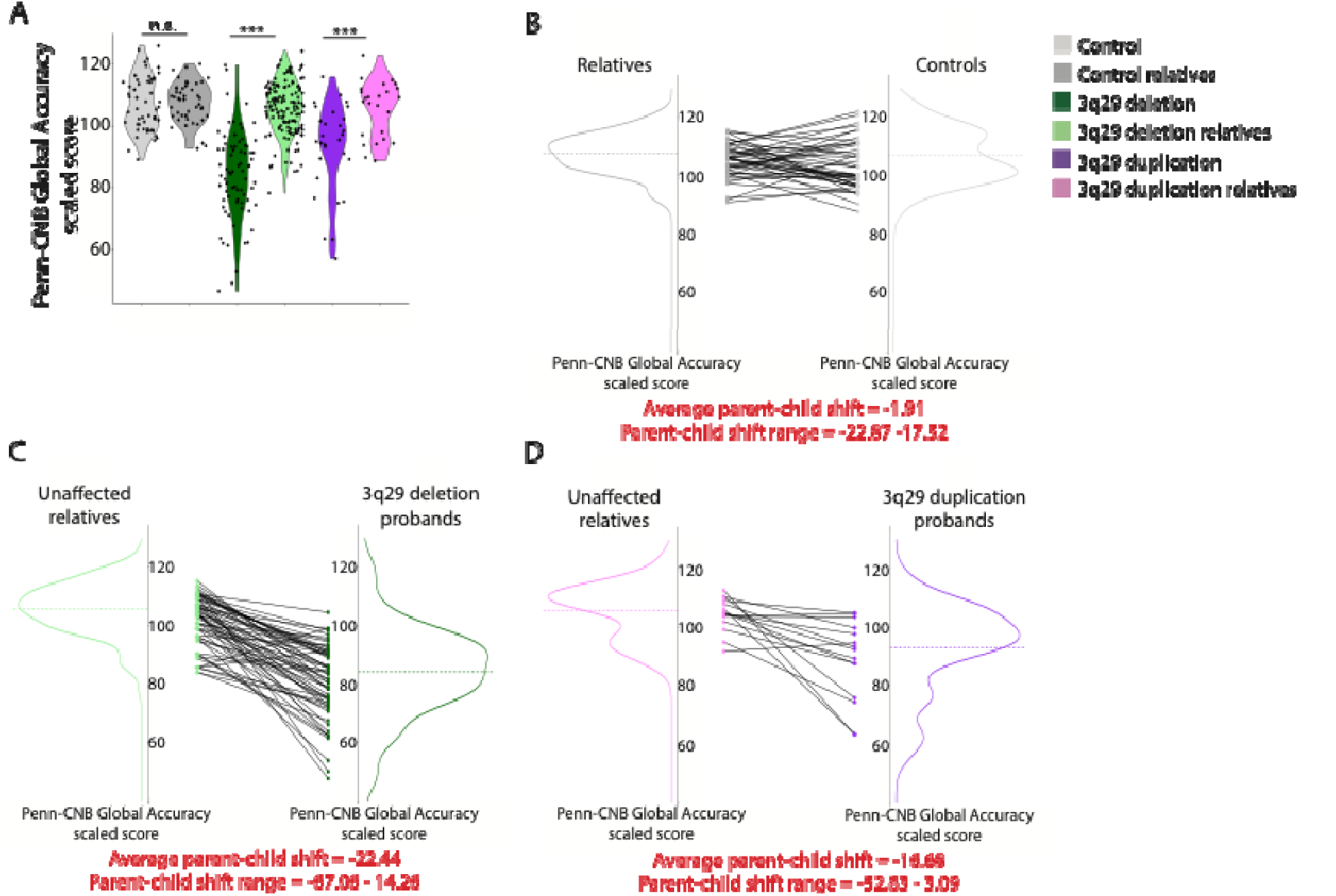
**A)** Distribution of Penn-CNB Global Accuracy scores for controls (n=54), control relatives (n=45), 3q29del probands (n=74), 3q29del relatives (n=139), 3q29dup probands (n=29), and 3q29dup relatives (n=25). **B)** Within-family relative-proband correlation for Penn-CNB Global Accuracy in controls. **C)** Within-family relative-proband correlation for Penn-CNB Global Accuracy in 3q29del. **D)** Within-family relative-proband correlation for Penn-CNB Global Accuracy in 3q29dup. n.s., not significant; ***, p<0.001

We compared the 3q29del and 3q29dup relatives to our typically developing controls to determine if there were any systematic differences in ascertainment of these samples. Unaffected relatives of subjects with 3q29del scored significantly lower on Penn-CNB Global Efficiency in comparison to typically developing controls (control mean=104.83±8.01, 3q29del relative mean=100.63±8.01; p=0.0008; Figure S3A), which was driven by a significantly slower response time in 3q29del relatives (control mean=99.87±8.05, 3q29del relative mean=95.13±8.70; p=0.0004; Figure S3C). There were no differences in Penn-CNB Global Accuracy between controls and any unaffected relative sample (all p>0.350; Figure S3B), suggesting that unaffected relatives of 3q29del and 3q29dup probands have similar cognitive ability compared to controls, but slowed neurocognitive processing speed.

#### Familial shift in Penn-CNB performance

We next examined the within-family effect of genetic background on the observed cognitive deficits in our 3q29del and 3q29dup subjects, by calculating the shift in Penn-CNB performance between the proband and their unaffected relative(s) within each family. In the phenotypically similar 22q11.2 and 16p11.2 reciprocal CNVs, familial background has been shown to contribute to the variability in expression of cognitive, behavioral, and physical phenotypes (48, 49, 50, 51, 52). There was no significant shift in Penn-CNB Global Accuracy between control subjects and their relatives (average shift=-1.91±9.83, p=0.199; Figure 4B). In contrast, we observed a significant negative shift in Penn-CNB Global Accuracy score associated with both 3q29del and 3q29dup. On average, subjects with 3q29del scored more than one standard deviation below their unaffected relatives on Global Accuracy (average shift=-18.87±12.31, p<2.2E-16), with shift values ranging from-56.87 to 11.98 (Figure 4C). Subjects with 3q29dup scored approximately one standard deviation below their unaffected relatives on Global Accuracy (average shift=-14.02±13.99, p=0.002), with shift values ranging from-44.33 to 2.60 (Figure 4D). The degree of shift in performance was significantly greater in both 3q29del (p=6.76E-12) and 3q29dup (p=0.0007) compared to controls. Sex and age of the proband did not have an impact on the degree of shift in cognition (all p>0.450). We repeated this analysis after removing any study subjects that did not have a biological first-degree relative as a comparator (n=0 control subjects (0% of proband-caregiver pairs), 2 3q29del subjects (3.33% of proband-caregiver pairs), and 3 3q29dup subjects (20% of proband-caregiver pairs)). Shift values were not different between the original and first-degree relative-only analyses (all p>0.750, Table S5), indicating that the inclusion of non-first-degree relatives did not influence the analysis. Together, these data indicate a significant negative shift in cognitive ability associated with 3q29del and 3q29dup.

### Relationship between CHR and cognition in 3q29del

It is well-established that the 3q29 deletion confers a greater than 40-fold increased risk for SZ (6, 7, 8, 9, 10). In the general population, individuals at Clinical High Risk (CHR) for psychosis exhibit impaired cognition (65, 66, 67, 68); cognitive function is also negatively associated with subthreshold psychosis symptoms in the phenotypically similar 22q11.2 deletion syndrome (69, 70). However, cognitive function in the context of CHR in individuals with 3q29del has not been explored. In our sample, subjects with 3q29del+CHR had significant impairments on the PPVT-5 relative to 3q29del-CHR (3q29del-CHR mean=93.33±12.53, 3q29del+CHR mean=81.76±14.19, p=9.53E-6; Figure 5A). On the Penn-CNB, subjects with 3q29del+CHR had a suggestive deficit on the Global Efficiency score (3q29del-CHR mean=87.05±14.11, 3q29del+CHR mean=80.62±16.46, p=0.05; Figure 5B) and the Global Accuracy score (3q29del-CHR mean=87.01±14.64, 3q29del+CHR mean=81.62±13.45, p=0.09; Figure 5C). There was no relationship between CHR status and the Penn-CNB Global Speed score (Figure 5D). These data highlight a pronounced deficit in verbal ability and subtle changes to neurocognition associated with CHR status in individuals with 3q29del.

**Figure 5.**
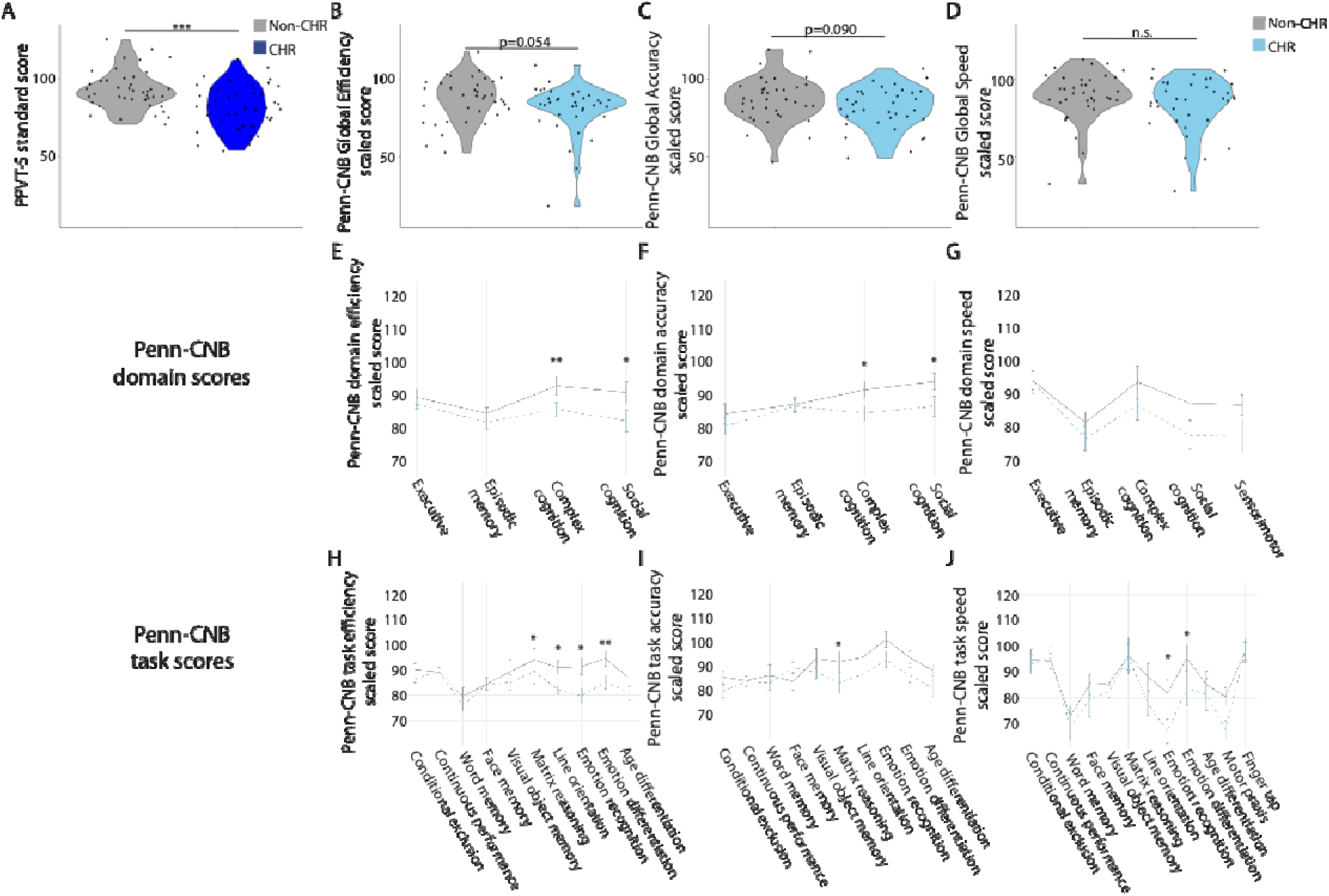
**A)** Distribution of PPVT-5 scores for non-CHR subjects with 3q29del (n=42) and CHR subjects with 3q29del (n=45). **B)** Distribution of Penn-CNB Global Efficiency scores for non-CHR subjects with 3q29del (n=37) and CHR subjects with 3q29del (n=36). **C)** Distribution of Penn-CNB Global Accuracy scores for non-CHR subjects with 3q29del (n=37) and CHR subjects with 3q29del (n=36). **D)** Distribution of Penn-CNB Global Speed scores for non-CHR subjects with 3q29del (n=37) and CHR subjects with 3q29del (n=36). **E)** Penn-CNB domain efficiency scores for non-CHR subjects with 3q29del (n=37) and CHR subjects with 3q29del (n=36). **F)** Penn-CNB domain accuracy scores for non-CHR subjects with 3q29del (n=37) and CHR subjects with 3q29del (n=36). **G)** Penn-CNB domain speed scores for non-CHR subjects with 3q29del (n=37) and CHR subjects with 3q29del (n=36). **H)** Penn-CNB task efficiency scores for non-CHR subjects with 3q29del (n=37) and CHR subjects with 3q29del (n=36). **I)** Penn-CNB task accuracy scores for non-CHR subjects with 3q29del (n=37) and CHR subjects with 3q29del (n=36). **J)** Penn-CNB task speed scores for non-CHR subjects with 3q29del (n=37) and CHR subjects with 3q29del (n=36). n.s., not significant; *, p<0.05; **, p<0.01; ***, p<0.001. Data are shown as mean ± SEM.

Studies of CHR individuals in the general population have identified differences in the neurocognitive profile associated with CHR status (65). To explore whether CHR and non-CHR individuals with 3q29del have different neurocognitive profiles, we examined the Penn-CNB domain and task scores. Individuals with 3q29del+CHR had significantly lower efficiency on the Complex cognition and Social cognition domains, with the largest deficit on the Complex cognition domain (3q29del-CHR mean=92.85 ±9.12, 3q29del+CHR mean=85.77±11.78, p=0.003; Figure 5E). These deficits were driven by significantly lower accuracy scores for 3q29del+CHR subjects in the Complex cognition and Social cognition domains, with the largest accuracy deficit on the Complex cognition domain (3q29del-CHR mean=91.67±15.74, 3q29del+CHR mean=84.57±11.88, p=0.03; Figure 5F). There was a suggestive association between CHR status and Social cognition response speed (3q29del-CHR mean=87.61±22.01, 3q29del+CHR mean=77.87±27.09, p=0.08; Figure 5G). At the task level, individuals with 3q29del+CHR had significantly worse efficiency scores on four of ten Penn-CNB tasks, with the worst deficit on the Emotion recognition task (3q29del-CHR mean=91.45±17.94, 3q29del+CHR mean=79.83±29.26, p=0.03; Figure 5H). Individuals with 3q29del+CHR had significantly lower accuracy scores on the Matrix reasoning task (3q29del-CHR mean=91.93±16.67, 3q29del+CHR mean=83.21±13.58, p=0.02; Figure 5I). Finally, individuals with 3q29del+CHR had significantly slower response speed on two of 12 Penn-CNB tasks, with the most substantial deficit on the Emotion differentiation task (3q29del-CHR mean=95.53 ±14.78, 3q29del+CHR mean=83.77±28.92, p=0.04; Figure 5J). Together, these data identify differences in neurocognition associated with CHR status in 3q29del, and potentially identify sentinel tasks that could be used as an early marker for psychosis risk.

## Discussion

The present study is the largest systematic evaluation of cognition in 3q29del and 3q29dup. Here we present a novel direct comparison of cognitive ability between probands with 3q29del and 3q29dup and their unaffected relatives. Both 3q29del and 3q29dup are associated with significant deficits in receptive verbal ability, measured by the PPVT-5, and across multiple cognitive domains, measured by the Penn-CNB, compared to unaffected controls. Additionally, subjects with 3q29del had significantly worse performance on the Penn-CNB than subjects with 3q29dup, on global metrics and on domain-and task-level scores. Both 3q29del and 3q29dup had significantly increased variability in Penn-CNB performance relative to controls and to their respective unaffected relatives, and both groups exhibit a significant negative shift in Penn-CNB performance when compared to their biological relatives. Finally, individuals with 3q29del+CHR have significantly worse performance on the PPVT-5 and on the Penn-CNB compared to individuals with 3q29del-CHR.

We identified specific differences in the neurocognitive profile between 3q29del and 3q29dup on efficiency, accuracy, and response speed. Studies of the phenotypically similar 22q11.2 and 16p11.2 reciprocal CNVs have also identified differences in the neurocognitive profile between subject groups (71, 72). In the present study, subjects with 3q29del had a more substantial impairment in accuracy in the Episodic memory domain, which is aligned with findings in 22q11.2 deletion and duplication syndromes (71). Prior work has shown that the 22q11.2 duplication is associated with slower response time relative to the 22q11.2 deletion (71, 72) and the 16p11.2 duplication is associated with slower response time relative to the 16p11.2 deletion (72); in the present study, we find that individuals with 3q29del record significantly slower response times on the Executive, Episodic memory, and Social cognition domains relative to 3q29dup. The pattern of decreased accuracy but increased response speed in 22q11.2 deletion syndrome (71, 73, 74) is consistent with studies of idiopathic ADHD (75) and is attributed to the concept of an accuracy/speed trade off. However, this does not appear to hold true in 3q29del; even though individuals with 3q29del are at increased risk for ADHD (17), our subjects with 3q29del exhibit both decreased accuracy and decreased response speed relative to controls and subjects with 3q29dup. These results suggest that the observed deficits in Penn-CNB performance in 3q29del are not related to attentional challenges but rather reflect underlying neurocognitive deficits in this population.

The present study is the first systematic comparison of cognitive ability between probands with 3q29del and 3q29dup and their unaffected relatives. We found that both 3q29del and 3q29dup are associated with a significant negative shift in the average Penn-CNB Global Accuracy score in comparison to the distribution of scores for their unaffected relatives. The Global Accuracy score distribution for subjects with 3q29del was shifted more than one standard deviation lower than the 3q29del biological relative distribution, and the distribution for subjects with 3q29dup was shifted one standard deviation lower than the 3q29dup relative distribution. The shift in performance in both 3q29del and 3q29dup was significantly larger than expected based on our typically developing controls, highlighting the adverse impact of the 3q29 CNVs on cognition. These data demonstrate that both 3q29 CNVs shift cognitive ability substantially below family baseline.

The progression of psychotic illness is linked with declines in cognitive ability; as the 3q29 deletion is an established SZ risk variant (6, 7, 8, 9, 10), we explored the relationship between subthreshold psychosis symptoms and cognition. Studies of CHR individuals in the general population have identified cognitive deficits associated with the presence and degree of subthreshold psychosis symptoms (65, 66, 67, 68), and cognitive impairment associated with subthreshold psychosis symptoms in 22q11.2 deletion syndrome (69, 70). The present study is the first to examine cognition in CHR individuals with 3q29del. We found a substantial deficit in verbal ability in individuals with 3q29del+CHR compared to 3q29del-CHR, which is consistent with prior work in 22q11.2 deletion syndrome (70). Further, we found differences in the neurocognitive profile between 3q29del+CHR and 3q29del-CHR individuals, with deficits in both Complex and Social cognition domains. Interestingly, subjects with 3q29del+CHR had deficits on both the Emotion recognition and Emotion differentiation tasks. Given that inappropriate and blunted affect are commonly present in individuals with SZ, it is possible that these tasks on the Penn-CNB may serve as early indicators of illness severity. Indeed, emotion recognition deficits have been identified in CHR individuals in the general population (76), and impairments in emotion recognition are associated with higher SZ polygenic risk score (77). Further work is needed to define the longitudinal predictive value of Penn-CNB tasks on the progression of psychotic illness in 3q29del.

While this is the largest systematic evaluation of cognition in 3q29del and 3q29dup, it is not without limitations. First, the PPVT-5 and Penn-CNB were deployed remotely in the present study, so we had limited ability to control the testing environment for our study subjects. However, the Penn-CNB has been validated for remote use in 22q11.2 deletion syndrome (78) and we have validated remote administration of both the PPVT-5 and Penn-CNB in 3q29del (53). Second, the average Penn-CNB Global Accuracy scores for both 3q29del and 3q29dup in the present study are higher than prior reports of cognitive ability in these populations (3, 4, 15, 16, 24). This is likely because the Penn-CNB requires basic verbal comprehension, focused attention, and motor skills; as such, the lowest-functioning individuals in our study population were unable to complete the assessment. Indeed, 50% of the 3q29del and 3q29dup subjects that were unable to complete the Penn-CNB were nonverbal, and the average IQ was below 70 for individuals unable to complete the Penn-CNB (3q29del non-completer n=26, 3q29del non-completer mean IQ=61.90±14.82; 3q29dup non-completer n=6, 3q29dup non-completer mean IQ=44.00±4.00; Table S1). This suggests that the cognitive ability for subjects in the present study may be an overestimate of the cognitive ability of the 3q29del and 3q29dup populations as a whole. Finally, we were underpowered in the present study to assess the influence of de novo vs inherited CNV status, 3q29 CNV size, and race and ethnicity on cognition. Future recruitment efforts targeting underrepresented populations, as well as targeted recruitment for inherited and non-canonical 3q29 CNVs, will help to address these outstanding questions.

This is the first systematic comparison of cognitive ability in 3q29del and 3q29dup, as well as the first comparison to unaffected relatives. We identified significant deficits in cognition in both 3q29del and 3q29dup, with more substantial deficits in 3q29del. Subjects with 3q29del had significantly decreased accuracy and significantly slower response time than subjects with 3q29dup, which is distinct from 22q11.2 deletion and duplication syndromes. We also found that within subjects with 3q29del, individuals with 3q29del+CHR exhibited greater cognitive deficits than individuals with 3q29del-CHR. Together, these data highlight the importance of cognitive evaluation for all individuals diagnosed with 3q29del and 3q29dup, to ensure access to interventions that will ultimately improve long-term outcomes.

## Data Availability

All data produced in the present study are available upon reasonable request to the authors.

## Acknowledgements

We gratefully acknowledge our study population, the 3q29 deletion and 3q29 duplication community, for their participation and commitment to research. We thank Dr. Raquel Gur, Dr. Ruben Gur, Dr. Tyler Moore, and Kosha Ruparel for their expert advice in the implementation and analysis of the Penn-CNB. This study was funded by the National Institute of Mental Health grant R01 MH126449 (PI Mulle).

## Author’s contributions

RMP performed the statistical analysis, produced all figures and tables, and wrote the manuscript. DVB, MKH, and JRP performed evaluations of all study participants. DVB, MKH, JRP, TI, RCG, REG, TMM, and KR helped with data interpretation. JGM edited the manuscript and provided guidance on analyzing and interpreting data. JGM was the principal investigator responsible for study direction. All authors participated in commenting on the drafts and have read and approved the final manuscript.

