## Supplemental Material for "The cognitive profile of the schizophrenia-associated 3q29 deletion syndrome and the reciprocal 3q29 duplication syndrome"

**Supplemental Methods**

### ***Penn Computerized Neurobehavioral Test Battery***

As a measure of cognitive ability, this study utilized the *Penn Computerized Neurocognitive Battery (CNB)*, which consists of a series of tasks designed to measure cognitive performance across multiple domains. In the present study, the *CNB* was conducted remotely; a certified researcher administers the battery with each participant over a face-to-face video Zoom session, providing instructions, ensuring task comprehension, and noting any potential problems that may affect data quality (i.e. distractions, poor internet connection, motor issues). The subtests included in the version of the battery utilized in the present study were selected based on the known profile of cognitive ability in 3q29 deletion. There were 13 subtests included in the battery, as follows:

1) Motor Praxis

2) Measured Emotion Differentiation Test

3) Facial Memory Test

BREAK POINT

4) Emotion Recognition Test

5) Word Memory Test

6) Age Differentiation Test

7) Short Line Orientation Test

BREAK POINT

8) Visual Object Learning Test

9) Matrix Analysis Test

10) Short Continuous Performance Test

BREAK POINT

11) Conditional Exclusion Test

12) Go/No Go Test

13) Short Computerized Finger Tapping Test

For analysis, CNB global accuracy score was computed as an average of the z-scored accuracy measurements for tasks 2-11 above. Note that for the Conditional Exclusion Test, the *acc2* measure was used as the accuracy score, which is (1+ number of categories achieved) x proportion of correct responses rather than the total number of correct responses because many correct responses can occur without solving a category. Motor Praxis and the Short Computerized Finger Tapping Test were not included in the global efficiency and global accuracy calculations, as these tasks do not have accuracy measurements. Age- and sex-normed z-scores were calculated based on population normative data available from the PennCNB (<https://webcnp.med.upenn.edu/cnb_reports/>). Normative data for subjects under 21 years of age was not available for the Go/No Go Task, so that task was excluded from analysis for all participants. For each task, participant completion and effort were scored on a scale of C1-C3 and I1-I3 (Figure S1). Tasks with a status code of C3, I1, I2, or I3 were removed from analysis, and the subject’s data for the other tasks was preserved. If a subject had a status code of C3, I1, I2, or I3 on 7 or more tasks (more than 50% of assessment), they were removed from the analysis. Tasks with a z-score less than -10 were considered extreme outliers and removed from analysis, and the subject’s data for the other tasks was preserved. Data were winsorized with a 1% threshold prior to analysis. Composite global accuracy score was based on the average of the tasks that pass QC for each participant, with no imputation for missing data.

**Supplemental Results**

In the present study, we were able to determine the precise genomic coordinates of the 3q29 CNV for 69 individuals with 3q29del (76.67% of total enrollment) and 18 individuals with 3q29dup (51.43% of total enrollment) via clinical genetics report review. For those individuals where we had access to a full clinical genetics report, 61 individuals with 3q29del had the canonical 1.6 Mb deletion (88.41% of available records), 3 individuals had a deletion larger than 1.6 Mb (4.35% of available records), and 5 individuals had a deletion smaller than 1.6 Mb (7.25% of available records). For subjects with 3q29dup, 7 individuals had the canonical 1.6 Mb duplication (38.89% of available records), 3 individuals had a duplication larger than 1.6 Mb (16.67% of available records), and 8 individuals had a duplication smaller than 1.6 Mb (44.44% of available records). We performed an exploratory analysis to determine if the observed heterogeneity in cognitive function in our subjects with 3q29del and 3q29dup was due to variability in the size of the CNV interval. CNV size had no association with PPVT-5 performance for subjects with 3q29del (all p>0.750, Figure S2A) or 3q29dup (all p>0.450, Figure S2E). CNV size had no association with Penn-CNB Global Accuracy for individuals with 3q29del or 3q29dup (all p>0.170, Figure S2C). We also determined inheritance patterns for study subjects with canonical 3q29del or 3q29dup. For the 61 subjects with canonical 3q29del, 55 cases were confirmed or presumed *de novo* (90.16% of canonical 3q29del) and 6 cases were confirmed inherited (9.84% of canonical 3q29del). For the 7 subjects with canonical 3q29dup, 4 cases were confirmed or presumed *de novo* (57.14% of canonical 3q29dup) and 3 cases were confirmed inherited (42.86% of canonical 3q29dup). Inheritance pattern did not have a significant effect on PPVT-5 performance or Penn-CNB Global Accuracy score for 3q29del or 3q29dup (all p>0.160, Figure S2B, D, F, H).

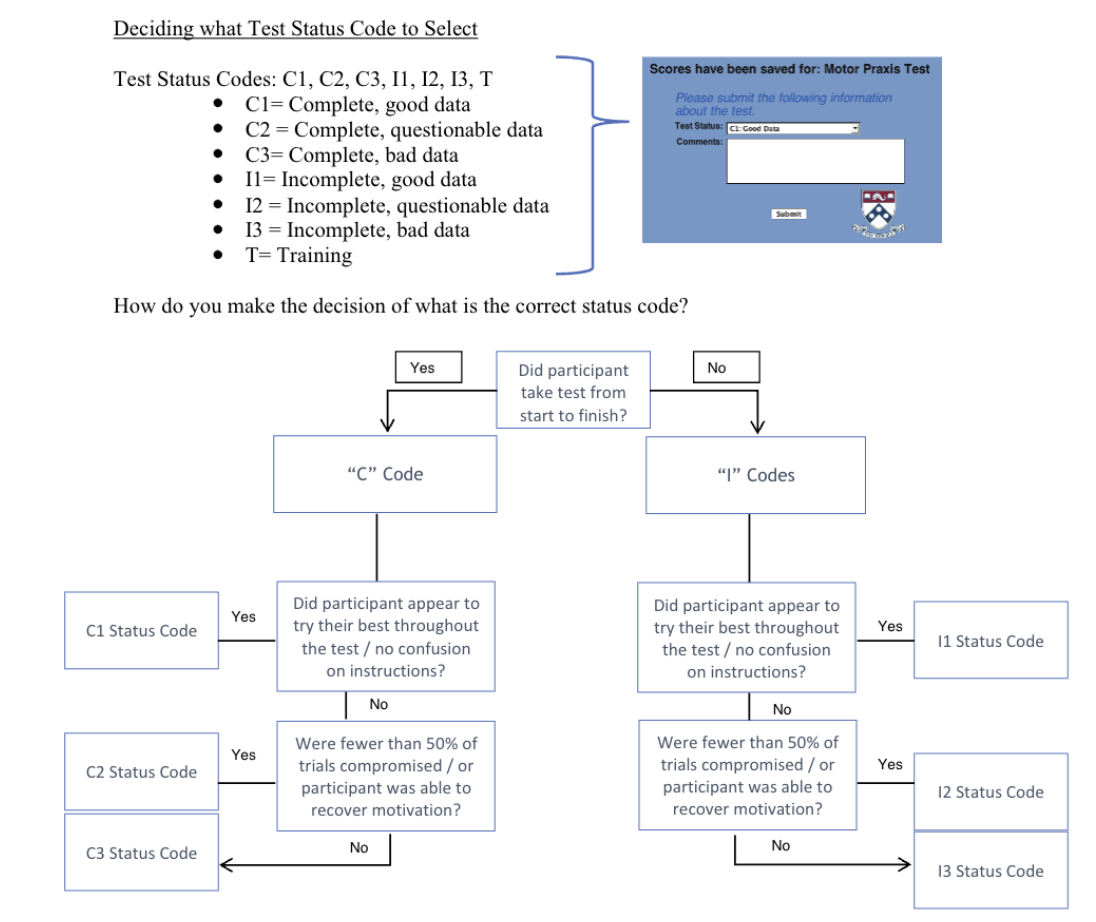

**Figure S1.** Decision tree illustrating Penn-CNB quality control process. Adapted from Penn-CNB manual.

**Table S1.** Demographics for study subjects that could not complete the Penn-CNB.

|  |  | 3q29 deletion | 3q29 duplication | 3q29 deletion caregivers |
| --- | --- | --- | --- | --- |
| N |  | 26 | 6 | 1 |
| Percent of total attempted Penn-CNBs |  | 25.74% | 17.14% | 0.66% |
| Age (years) |  |  |  |  |
|  | Mean ± SD | 11.42 ± 4.31 | 10.17 ± 4.45 | 52 |
|  | Range | 7 - 24 | 7 - 19 |  |
| External Composite IQ |  |  |  |  |
|  | Other IQ testing available | 16 (61.54%) | 3 (50.00%) | 0 (0.00%) |
|  | No other IQ testing available | 10 (38.46%) | 3 (50.00%) | 1 (100.00%) |
|  | Mean ± SD | 61.90 ± 14.82 | 44.00 ± 4.00 |  |
|  | Range | 40 - 84 | 40 - 48 |  |
| Sex |  |  |  |  |
|  | Male | 20 (76.92%) | 3 (50.00%) | 1 (100.00%) |
|  | Female | 6 (23.08%) | 3 (50.00%) | 0 (0.00%) |
| Race |  |  |  |  |
|  | White | 25 (96.15%) | 5 (83.33%) | 1 (100.00%) |
|  | Other | 1 (3.85%) | 1 (16.67%) | 0 (0.00%) |
| Ethnicity |  |  |  |  |
|  | Hispanic/Latino | 1 (3.85%) | 2 (33.33%) | 0 (0.00%) |
|  | Not Hispanic/Latino | 25 (96.15%) | 4 (66.67%) | 1 (100.00%) |
| Reason for not completing Penn-CNB |  |  |  |  |
|  | Nonverbal | 13 (50.00%) | 3 (50.00%) | 0 (0.00%) |
|  | Unable to complete | 0 (0.00%) | 1 (16.67%) | 0 (0.00%) |
|  | Failed more than 50% of tasks | 12 (46.15%) | 2 (33.33%) | 0 (0.00%) |
|  | Extreme outliers on more than 50% of tasks | 1 (3.85%) | 0 (0.00%) | 1 (3.85%) |

**Table S2.** Penn-CNB domain and task efficiency scores.

|  |  | Control | 3q29 deletion | | 3q29 duplication | |  |
| --- | --- | --- | --- | --- | --- | --- | --- |
|  |  | Mean ± SD | Mean ± SD | p value vs Control | Mean ± SD | p value vs Control | p value, 3q29 deletion vs 3q29 duplication |
| Domains | Executive | 102.99 ± 8.86 | 88.66 ± 13.17 | **4.61E-11** | 96.14 ± 9.75 | **0.011** | **0.003** |
|  | Episodic memory | 100.54 ± 7.72 | 83.40 ± 18.21 | **8.40E-10** | 91.87 ± 14.03 | **0.011** | **0.014** |
|  | Complex cognition | 103.73 ± 6.49 | 89.28 ± 10.97 | **3.26E-14** | 94.55 ± 10.62 | **6.25E-05** | **0.032** |
|  | Social cognition | 105.08 ± 9.41 | 86.49 ± 16.60 | **2.56E-12** | 96.33 ± 11.76 | **0.006** | **0.001** |
| Tasks | Conditional exclusion | 107.28 ± 9.59 | 88.43 ± 13.01 | **8.79E-16** | 94.25 ± 10.25 | **3.46E-06** | **0.024** |
|  | Continuous performance | 99.29 ± 12.21 | 89.82 ± 18.50 | **0.001** | 98.04 ± 13.35 | 0.744 | **0.024** |
|  | Word memory | 100.04 ± 12.60 | 77.93 ± 28.08 | **1.11E-07** | 94.77 ± 16.49 | 0.3 | **0.0006** |
|  | Face memory | 101.68 ± 9.76 | 84.61 ± 18.58 | **1.19E-08** | 92.03 ± 16.81 | **0.011** | 0.067 |
|  | Visual object memory | 99.89 ± 8.69 | 87.30 ± 17.98 | **3.67E-06** | 90.54 ± 13.55 | **0.006** | 0.33 |
|  | Matrix reasoning | 100.64 ± 7.78 | 91.86 ± 9.80 | **5.72E-07** | 94.37 ± 10.52 | **0.005** | 0.289 |
|  | Line orientation | 106.82 ± 9.08 | 86.46 ± 15.10 | **5.40E-15** | 96.50 ± 11.61 | **0.0008** | **0.001** |
|  | Emotion recognition | 107.32 ± 13.74 | 85.74 ± 24.55 | **6.92E-09** | 96.32 ± 13.63 | **0.016** | **0.007** |
|  | Emotion differentiation | 106.16 ± 9.53 | 89.63 ± 15.20 | **8.85E-11** | 97.15 ± 13.88 | **0.004** | **0.019** |
|  | Age differentiation | 101.77 ± 10.81 | 83.99 ± 17.79 | **5.58E-10** | 95.51 ± 13.55 | 0.07 | **0.0008** |

**Table S3.** Penn-CNB domain and task accuracy scores.

|  |  | Control | 3q29 deletion | | 3q29 duplication | |  |
| --- | --- | --- | --- | --- | --- | --- | --- |
|  |  | Mean ± SD | Mean ± SD | p value vs Control | Mean ± SD | p value vs Control | p value, 3q29 deletion vs 3q29 duplication |
| Domains | Executive | 102.94 ± 11.72 | 82.58 ± 15.85 | **7.03E-14** | 89.80 ± 11.02 | **7.94E-05** | **0.013** |
|  | Episodic memory | 102.04 ± 9.94 | 86.73 ± 15.18 | **7.41E-09** | 90.76 ± 16.75 | **0.0006** | 0.267 |
|  | Complex cognition | 111.37 ± 10.19 | 87.93 ± 14.38 | **<2E-16** | 96.60 ± 11.37 | **1.19E-06** | **0.002** |
|  | Social cognition | 107.25 ± 9.45 | 90.07 ± 17.04 | **1.24E-09** | 99.46 ± 16.86 | **0.024** | **0.014** |
| Tasks | Conditional exclusion | 106.83 ± 14.16 | 82.41 ± 15.01 | **5.14E-16** | 89.59 ± 14.60 | **1.77E-06** | **0.037** |
|  | Continuous performance | 99.55 ± 17.87 | 84.20 ± 24.52 | **0.0001** | 91.18 ± 16.35 | 0.107 | 0.124 |
|  | Word memory | 99.98 ± 14.68 | 84.89 ± 25.18 | **0.0003** | 92.70 ± 26.44 | 0.166 | 0.197 |
|  | Face memory | 103.89 ± 13.74 | 86.32 ± 17.00 | **7.60E-09** | 91.99 ± 16.75 | **0.002** | 0.148 |
|  | Visual object memory | 102.23 ± 13.05 | 90.51 ± 16.23 | **2.16E-05** | 91.39 ± 14.35 | **0.002** | 0.791 |
|  | Matrix reasoning | 108.60 ± 13.10 | 87.70 ± 15.76 | **2.21E-12** | 94.97 ± 16.81 | **0.0002** | 0.059 |
|  | Line orientation | 114.13 ± 11.88 | 89.86 ± 16.06 | **<2E-16** | 99.96 ± 9.91 | **3.26E-05** | **0.0007** |
|  | Emotion recognition | 112.48 ± 13.77 | 96.73 ± 21.86 | **9.87E-06** | 102.60 ± 20.38 | **0.026** | 0.205 |
|  | Emotion differentiation | 106.56 ± 12.50 | 89.58 ± 18.35 | **7.98E-08** | 99.18 ± 19.59 | 0.059 | **0.027** |
|  | Age differentiation | 102.71 ± 15.73 | 84.79 ± 20.78 | **4.85E-07** | 96.59 ± 19.57 | 0.162 | **0.009** |

**Table S4.** Penn-CNB domain and task speed scores.

|  |  | Control | 3q29 deletion | | 3q29 duplication | |  |
| --- | --- | --- | --- | --- | --- | --- | --- |
|  |  | Mean ± SD | Mean ± SD | p value vs Control | Mean ± SD | p value vs Control | p value, 3q29 deletion vs 3q29 duplication |
| Domains | Executive | 103.04 ± 11.02 | 94.73 ± 18.27 | **0.003** | 102.47 ± 12.46 | 0.874 | **0.019** |
|  | Episodic memory | 99.04 ± 12.38 | 80.07 ± 30.50 | **2.62E-05** | 92.98 ± 23.74 | 0.282 | **0.026** |
|  | Complex cognition | 96.09 ± 12.22 | 90.64 ± 20.02 | 0.078 | 92.50 ± 16.88 | 0.368 | 0.641 |
|  | Social cognition | 102.92 ± 14.00 | 82.92 ± 24.81 | **1.45E-07** | 93.20 ± 16.88 | **0.039** | **0.018** |
|  | Sensorimotor | 103.30 ± 12.40 | 82.86 ± 28.48 | **3.84E-06** | 88.88 ± 26.81 | **0.009** | 0.319 |
| Tasks | Conditional exclusion | 107.72 ± 10.39 | 94.45 ± 20.48 | **2.01E-05** | 98.91 ± 14.61 | **0.025** | 0.237 |
|  | Continuous performance | 99.03 ± 15.79 | 95.43 ± 23.85 | 0.329 | 104.89 ± 17.01 | 0.237 | **0.041** |
|  | Word memory | 100.10 ± 17.60 | 70.97 ± 43.63 | **2.10E-06** | 96.84 ± 17.64 | 0.666 | **0.0001** |
|  | Face memory | 99.48 ± 13.71 | 82.90 ± 31.86 | **0.0006** | 92.07 ± 27.90 | 0.236 | 0.174 |
|  | Visual object memory | 97.53 ± 13.31 | 84.09 ± 30.83 | **0.003** | 89.69 ± 25.27 | 0.174 | 0.351 |
|  | Matrix reasoning | 92.68 ± 18.25 | 96.03 ± 20.72 | 0.356 | 93.78 ± 21.40 | 0.815 | 0.642 |
|  | Line orientation | 99.51 ± 14.11 | 83.05 ± 25.47 | **2.65E-05** | 93.03 ± 17.24 | 0.192 | **0.038** |
|  | Emotion recognition | 102.17 ± 18.60 | 74.75 ± 36.24 | **4.71E-07** | 90.05 ± 23.66 | 0.071 | **0.015** |
|  | Emotion differentiation | 105.75 ± 14.80 | 89.68 ± 23.31 | **9.55E-06** | 95.12 ± 16.85 | **0.02** | 0.193 |
|  | Age differentiation | 100.84 ± 15.29 | 83.18 ± 28.34 | **4.96E-05** | 94.43 ± 22.68 | 0.238 | **0.04** |
|  | Motor praxis | 99.03 ± 19.10 | 74.40 ± 38.54 | **3.65E-05** | 84.00 ± 34.34 | **0.045** | 0.224 |
|  | Finger tap | 113.97 ± 17.35 | 98.25 ± 24.11 | **2.40E-05** | 103.03 ± 12.29 | **0.02** | 0.189 |

**Table S5.** Comparison of familial shift analysis in full dataset and first-degree relatives only dataset.

|  | Control | | | 3q29 deletion | | | 3q29 duplication | | |
| --- | --- | --- | --- | --- | --- | --- | --- | --- | --- |
|  | All biological relatives | First degree relatives only | p value | All biological relatives | First degree relatives only | p value | All biological relatives | First degree relatives only | p value |
| Caregiver-proband pairs (n) | 45 | 45 |  | 60 | 58 |  | 15 | 12 |  |
| Caregiver Penn-CNB Global Accuracy (mean ± SD) | 107.94 ± 6.32 | 107.94 ± 6.32 |  | 104.42 ± 9.20 | 104.54 ± 9.19 |  | 107.29 ± 6.17 | 108.96 ± 4.22 |  |
| Proband Penn-CNB Global Accuracy (mean ± SD) | 106.03 ± 8.99 | 106.03 ± 8.99 |  | 81.98 ± 13.55 | 82.16 ± 13.47 |  | 90.60 ± 14.79 | 90.98 ± 15.83 |  |
| Caregiver - child shift (mean ± SD) | -1.91 ± 9.83 | -1.91 ± 9.83 | 1 | -22.44 ± 14.63 | -22.38 ± 14.84 | 0.983 | -16.68 ± 16.66 | -17.98 ± 17.69 | 0.848 |
| Caregiver - child shift range | -22.87 - 17.52 | -22.87 - 17.52 |  | -67.06 - 14.26 | -67.06 - 14.26 |  | -52.83 - 3.09 | -52.83 - 1.04 |  |

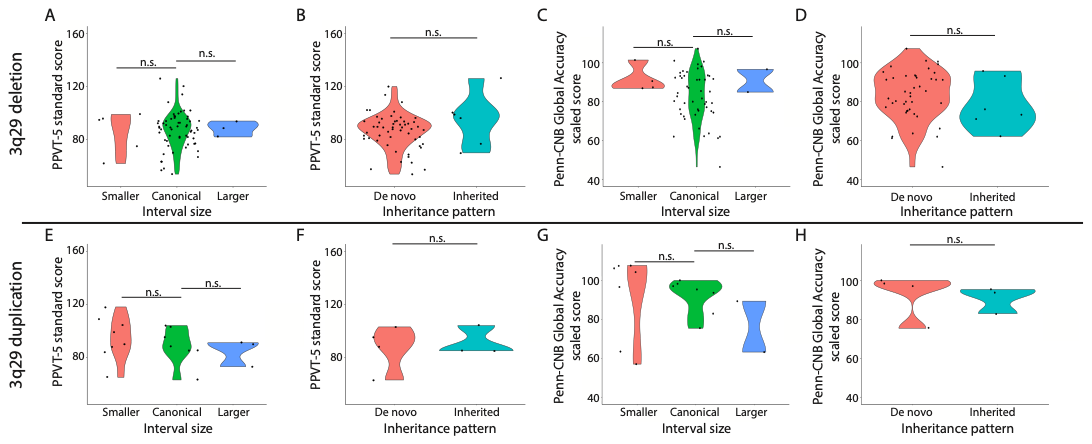

**Figure S2. A)** Distribution of PPVT-5 scores by CNV size for 3q29del. **B)** Distribution of PPVT-5 scores by inheritance status for 3q29del. **C)** Distribution of Penn-CNB Global Accuracy scores by CNV size for 3q29del. **D)** Distribution of Penn-CNB Global Accuracy scores by inheritance status for 3q29del. **E)** Distribution of PPVT-5 scores by CNV size for 3q29dup. **F)** Distribution of PPVT-5 scores by inheritance status for 3q29dup. **G)** Distribution of Penn-CNB Global Accuracy scores by CNV size for 3q29dup. **H)** Distribution of Penn-CNB Global Accuracy scores by inheritance status for 3q29dup.

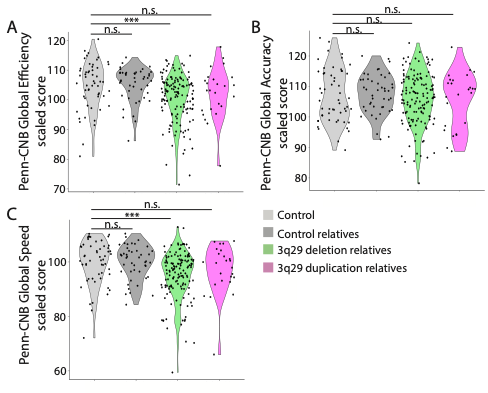

**Figure S3. A)** Distribution of Penn-CNB Global Efficiency scores for controls (n=54), control relatives (n=45), 3q29del relatives (n=139), and 3q29dup relatives (n=25). **B)** Distribution of Penn-CNB Global Accuracy scores for controls (n=54), control relatives (n=45), 3q29del relatives (n=139), and 3q29dup relatives (n=25). **C)** Distribution of Penn-CNB Global Speed scores for controls (n=54), control relatives (n=45), 3q29del relatives (n=139), and 3q29dup relatives (n=25). n.s., not significant; *, p<0.05; **, p<0.01; ***, p<0.001
